# Mapping obesity-related traits with regional white matter microstructure highlights the importance of brainstem tracts and sex-related differences

**DOI:** 10.1101/2025.01.24.25321078

**Authors:** Tiril P. Gurholt, Dani Beck, Irene Voldsbekk, Nadine Parker, Daniel E. Askeland-Gjerde, Ann-Marie G. de Lange, Dennis van der Meer, Christian K. Tamnes, Paul M. Thompson, Ida E. Sønderby, Ivan I. Maximov, Lars T. Westlye, Ole A. Andreassen

**Affiliations:** Section for Precision Psychiatry, Division of Mental Health and Addiction, Oslo University Hospital, Oslo, Norway; Department of Psychology, University of Oslo, Oslo, Norway; Division of Mental Health and Substance Abuse, Diakonhjemmet Hospital, Oslo, Norway; PROMENTA Research Center, Department of Psychology, University of Oslo, Oslo, Norway; Centre for Precision Psychiatry, Division of Mental Health and Addiction, Institute of Clinical Medicine, University of Oslo, Oslo, Norway; Institute of Clinical Medicine, University of Oslo, Oslo, Norway; Department of Clinical Neurosciences, Lausanne University Hospital (CHUV) and University of Lausanne, Lausanne, Switzerland; Department of Psychiatry, University of Oxford, Oxford, UK; School of Mental Health and Neuroscience, Faculty of Health, Medicine and Life Sciences, Maastricht University, Maastricht, The Netherlands; Imaging Genetics Center, Mark & Mary Stevens Neuroimaging & Informatics Institute, Keck School of Medicine, University of Southern California, Marina del Rey, CA, USA; Department of Medical Genetics, Oslo University Hospital, Oslo, Norway; K.G. Jebsen Centre for Neurodevelopmental Disorders, University of Oslo, Oslo, Norway; Department of Health and Functioning, Western Norway University of Applied Sciences, Bergen, Norway

**Keywords:** body mass index, waist-to-hip ratio, waist circumference, brain MRI, mental, psychiatric, neurological, somatic, disease, disorder

## Abstract

**Background and aims:** The relationships between obesity-related traits and the brain’s white matter characteristics and the context of sex- and age-related differences, remain unclear. This study aims to elucidate these body-brain connections using a large-scale dataset.

**Methods:** We analyzed data from 40,040 participants from the UK Biobank (52.2% female; ages 44-83 years) using multiple linear regression to evaluate associations between obesity-related traits (obesity, body anthropometrics) and white matter diffusion tensor imaging (DTI) metrics (fractional anisotropy, axial diffusivity, radial diffusivity, mean diffusivity). We also examined interactions with age and sex.

**Results:** Our analyses revealed significant associations between obesity-related traits and DTI metrics with partial correlation coefficient |r| effect sizes ranging from 0.02 to 0.20 for most regions of interest with largest effects in brainstem tracts. We observed more widespread sex-by-obesity-related than age-by-obesity-related interaction effects on DTI metrics.

**Conclusions:** Our results link obesity-related traits and white matter phenotypes, suggesting that shared body fat-related pathways linking physical and brain health that may vary based on sex and age. Understanding these relationships could enhance the development and evaluation of targeted, individualized, treatment strategies for conditions that co-occur with obesity, although further longitudinal studies are needed to map the dynamics of these associations.

## Introduction

Obesity and its associated cardiometabolic conditions are frequently observed alongside neurological and psychiatric brain disorders^1–3^. Research has shown that both obesity-related traits^4–9^ and brain disorders^10^ are linked to brain white matter alterations, possibly due to common underlying mechanisms. There are also notable sex- and age-related variations in body fat distribution^11^, brain white matter magnetic resonance imaging (MRI) phenotypes^12^, and the prevalence and clinical features of common brain disorders^3,13,14^. Therefore, gaining a deeper understanding of how obesity-related traits relate to white matter phenotypes, and whether and how these relationships are influenced by age and sex, is crucial for unraveling the complex comorbidities between cardiometabolic conditions and brain disorders.

Diffusion magnetic resonance imaging (dMRI) is a widely used technique for non-invasive, *in vivo,* characterization and quantification of white matter organization and microstructure. Diffusion tensor imaging (DTI)^15^ is sensitive to the direction and magnitude of water molecule diffusion in brain tissue^16^. From the diffusion tensor, various diffusion measures can be derived, including fractional anisotropy (FA; the degree of rotation invariant anisotropic diffusion), axial diffusivity (AD; the diffusivity along the principal eigenvector (i.e., main diffusion direction)), radial diffusivity (RD; the diffusivity perpendicular to the principal eigenvector (i.e., non-main diffusion)), and mean diffusivity (MD; the average diffusivity over eigenvectors)^16^. These measures provide insight into the underlying white matter tissue properties. Interpretation can be challenging, but lower FA is often observed in disorders and interpreted as not favorable, while lower AD, higher RD, and higher MD may indicate axonal degeneration, demyelination, and inflammation, respectively^16^. When analyzed together, these DTI metrics provide deeper insights into tissue properties and of regional variation that may be relevant for disentangling disease processes.

Prior studies suggest that obesity (i.e., body mass index (BMI)≥30) and other obesity-related traits (e.g., BMI, waist-to-hip ratio (WHR), and waist circumference) are associated with dMRI white matter phenotypes. The most consistent findings include widespread lower FA at higher BMI and WHR^4–8^, with indications of higher FA for some brainstem tracts^4^. A meta-analysis reported lower FA in the genu of the corpus callosum in obesity^9^. For AD and RD, prior reports are mixed^6^. Few larger studies exist^4,7–9,17,18^, and they differ in focus, investigated regions, and methods. They studied obesity^9^ or other obesity-related traits (BMI^4,7,8^; WHR^4,8^; total body fat percentage^17^; muscle fat infiltration^18^). Most reported on FA^4,7–9,17,18^, two reported on MD^4,17^, and none reported on AD and RD. There is a need for large-scale studies that investigate obesity and other obesity-related traits with regional white matter characteristics assessed using different DTI metrics to obtain deeper insight into the link between obesity-related traits and brain tissue properties.

Sex- and age-related differences in body shape and fat distribution^11^ and dMRI features^19–21^ might contribute to the reported links between obesity and white matter phenotypes. A review reported negative correlations between BMI and FA in several brain regions and indications of accelerated white matter ageing in obese individuals^5^. Higher white matter brain age has been associated with higher body fat^22,23^ and other cardiometabolic risk factors^24,25^, and one study indicated multisystem aging with both shared and unique body and brain aging processes^26^. Further, interactions between body fat and sex may influence white matter microstructure^17^, with a stronger association between higher white matter brain age and body fat in males than females^23^, possibly reflecting that the normative range of body fat is higher for females than males^27^.

To enhance our understanding of the links between physical and brain health and of putative shared obesity-related pathways, in the largest study to date, we investigated the links between obesity-related traits (i.e., obesity, BMI, WHR, and waist circumference) and DTI-based brain white matter phenotypes among 40,040 middle-aged to older adults (52.2% female) from the UK Biobank (https://www.ukbiobank.ac.uk). To target putative complex interplays with sex and age, we also tested for interactions between obesity-related traits and both sex and age on white matter phenotypes (**Figure 1**).

**Figure 1:**
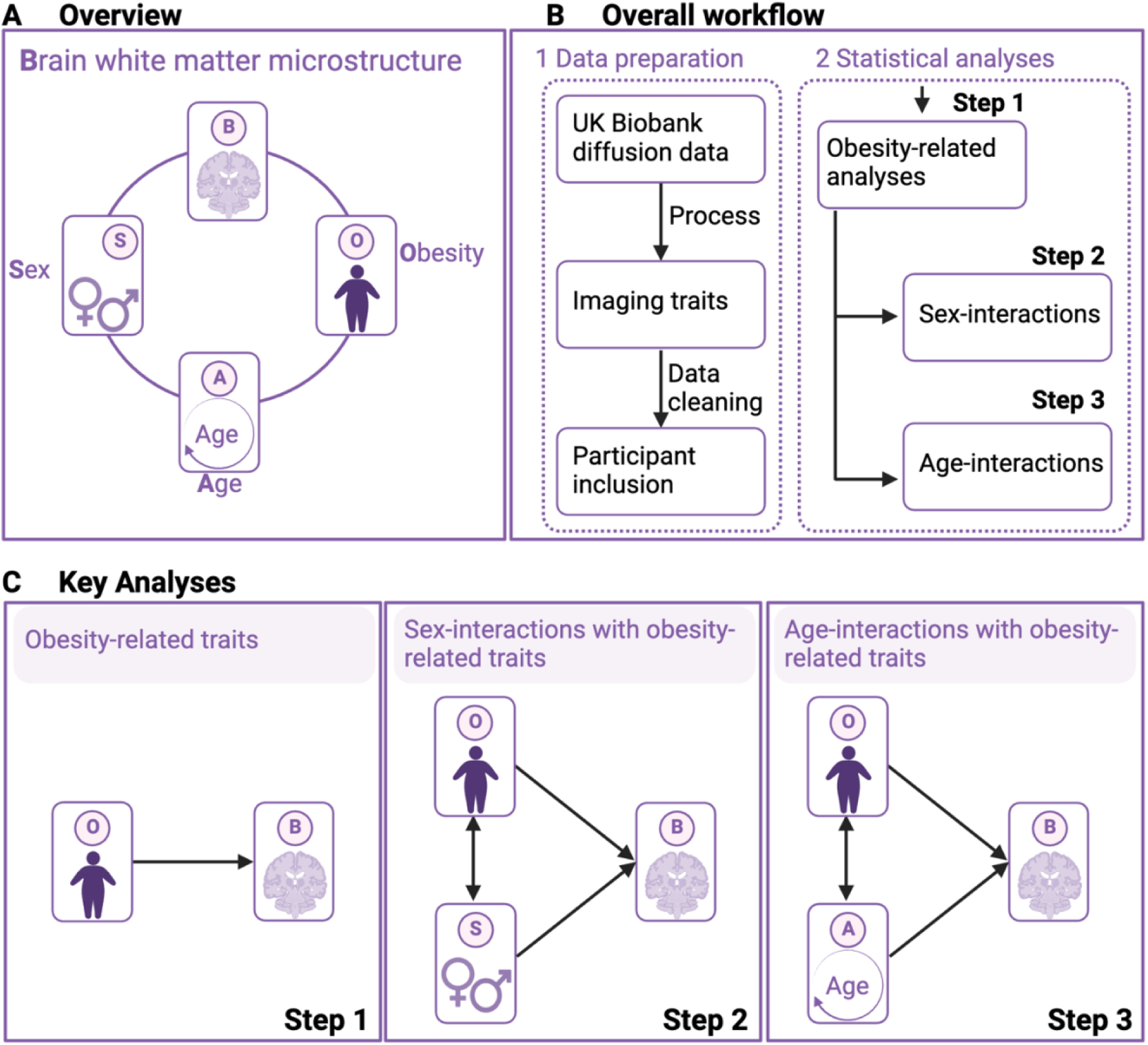
Overview of study design and analyses. (A) Overview of the study design targeting age, sex, and obesity-related traits relative to white matter microstructure. Description of the (B) study workflow and (C) key analyses (Created with BioRender.com).

## Methods

### Study Design and Participants

The UK Biobank is an epidemiological longitudinal population cohort of middle- to old-aged participants recruited from assessment sites across the UK^28^. The baseline assessment took place during 2006-2010 and included approximately 500K participants. The follow-up brain MRI assessment started in 2014 and is ongoing. We included 40,040 participants (20,904 females, 19,136 males) with brain dMRI and demographic and clinical data. We excluded participants who withdrew their informed consent (opt-out-list dated April 25^th^, 2023).

The UK Biobank has IRB approval from the North-West Multi-center Research Ethics Committee and obtained informed consent from all participants^28^. We obtained access to the UK Biobank resources through Application number 27412. We have approval from the Norwegian Regional Committees for Medical and Health Research Ethics (https://rekportalen.no/).

### Demographic and Clinical Data

**Table S1** summarizes the demographic and clinical data. We extracted age, sex, body anthropometrics (waist circumference, hip circumference, weight, height, BMI), and self-reported ethnicity, history of diabetes, hypercholesterolemia, hypertension, cigarette smoking, and alcohol consumption (**Note S1**). We computed waist-to-hip ratio (WHR), and derived binary (yes/no) variables for obesity (BMI≥30) and self-reported European/non-European ethnicity (i.e., white, white British, white Irish, other white background vs. other background), current cigarette smoking, and current alcohol consumption. We complemented self-reported ethnicity at MRI with data from the baseline assessment when necessary.

### MRI Acquisition and post-processing

MRI acquisition details are described elsewhere^28,29^. Briefly, 3D multimodal MRI of the brain was performed at four separate sites (Cheadle, Reading, Newcastle, or Bristol) in the UK, with similar 3T Siemens Skyra scanners, Siemens 32-channel head coils, and scanning protocols across sites.

We post-processed the available dMRI data using an optimized pipeline^30^ (**Note S2**). We derived DTI scalar metrics using a cumulant expansion of the diffusion signal^30^ with MATLAB scripts (https://github.com/NYU-DiffusionMRI/DESIGNER-v1). We normalized DTI metrics using FSL tract-based spatial statistics (TBSS)^31^ and harmonized them using the YTTRIUM algorithm^32^. All DTI metrics were skeletonized for the following averaging, extraction and tabularization. We extracted the 27 regions of interest (ROIs) for each diffusion map from the Johns Hopkins University (JHU) DTI atlas^33^, subsequently partitioned into brainstem, projection, commissural, and association pathways (**Figure 2**).

**Figure 2:**
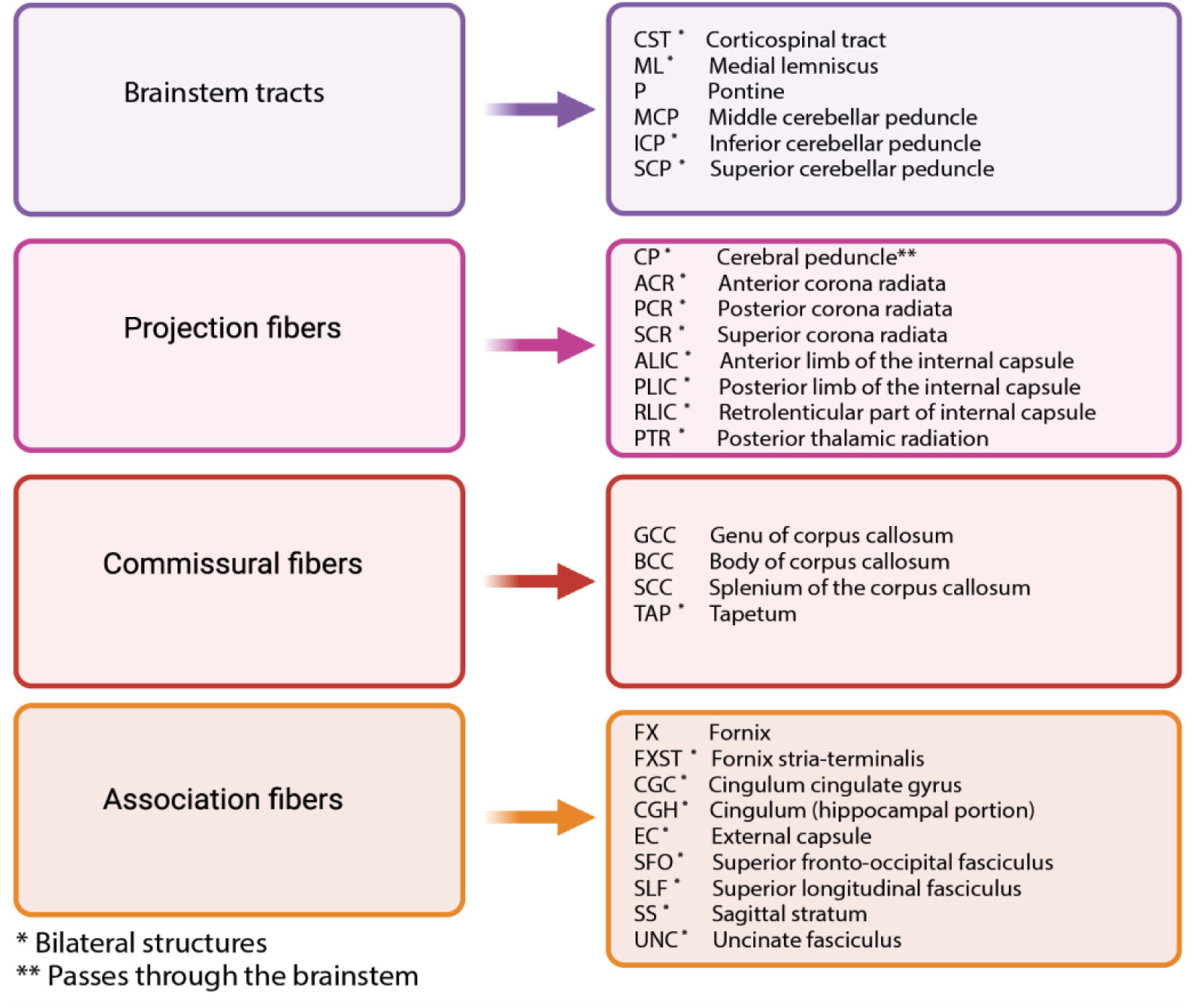
Overview of the included white matter regions of interest (ROIs) (Created with BioRender.com).

### Quality Control

Based on YTTRIUM for automatic dMRI quality control^32^, we removed 670 participants with poor dMRI data quality. We also removed five participants with incomplete diffusion data. In addition, we removed 1,839 participants with incomplete demographic or clinical data, resulting in a final sample of 40,040 participants (**Figure S1**).

### Statistical Analysis

We investigated the sample demographic and clinical data across and within sexes and for obesity vs. non-obesity (**Table S1**). We compared categorical variables using the χ^2^-test. We evaluated continuous variables for normality by visual inspection of density plots (**Figure S2**) and compared normally distributed variables using the two-sample t-test/Welch approximation for equal/unequal variance. We assessed density plots of diffusion metrics for expected distribution patterns (data not shown; all reasonably normal).

Initially (**Step 1)**, for each ROI and DTI metric, we tested for associations with obesity-related traits (i.e., obesity vs. non-obesity, BMI, WHR, and waist circumference) using linear models, while covarying for age, age^2^, sex, age-by-sex, age^2^-by-sex, ethnicity, and assessment site. Subsequently (**Step 2**), we included obesity-related trait-by-sex interactions while covarying for the corresponding main effects, age, age^2^, ethnicity, and assessment site. Lastly (**Step 3**), we included obesity-related trait-by-age interactions while covarying for the corresponding main effects, sex, ethnicity, and assessment site.

We implemented all statistical analyses in R (version 4.2.1; https://www.r-project.org). We adjusted all models for self-reported ethnicity since ethnicity is considered a relevant factor for body fat accumulation and obesity assessment^11^. We mean-centered all continuous regressors and entered binary variables as factors. We computed the partial correlation coefficient *r*-effect size directly from the t-statistics for continuous variables and via Cohen’s *d* for categorical variables^34^. We implemented a study-wide Bonferroni multiple comparison correction at α=0.05 across N=27×6×4=648 independent tests, reflecting the 27 ROIs, 6 main variables of interest (i.e., obesity vs. non-obesity, BMI, WHR, and waist circumference, and sex- and age-interactions), and 4 DTI metrics, counting partly overlapping models once, yielding a study-wide significance threshold of *p*≤α/N≈7.7e-05. We focused on the overall patterns and the most significant findings and reported the range of *p*-values and *r-effect* sizes (absolute values are indicated by |*r*|). Some p-values are rounded to zero since they are approximately zero and below numeric precision (i.e., smaller value than can be represented). We report the full results in the supplemental material.

## Results

### Demographic and clinical data

**Table S1** presents the demographic and clinical data. Briefly, the sample included more females (n=20,904; 52.2%; mean age 63.7±7.6 years) than males (n=19,136; 47.8%; mean age 64.9±7.8 years), had age range 44-83 years, and consisted predominantly of self-identified white Europeans (97.0%). On average, males exhibited higher levels of anthropometric traits for all measures (except hip circumference) compared to females. Participants with obesity (N=6,912; 50.7% female) had, on average, lower age, higher body anthropometric traits (except lower height), fewer alcohol consumers, and higher prevalence of self-reported cardiometabolic diagnosis than non-obese participants (N=33,192; 52.5% female).

### Associations between obesity-related traits and DTI metrics

Multiple linear regression analyses (**Step 1**) indicated similar associations with DTI metrics for both obesity and BMI (**Figure 3**; **Tables S2-S3**), albeit with generally stronger effects for the continuous BMI (|*r*| in [0.02, 0.20], p in [5.5e-05, 0〉 (i.e., p-value range from 5.5e-05 to approximately 0)) than the categorical obesity measure (|*r*| in [0.02, 0.14], p in [6.6e-05, 8.1e-185]). The number of significant ROIs was slightly larger for BMI (79.2%) than for obesity (76.6%), and largest for FA (obesity: 87.5%; BMI: 87.5%) followed by MD (obesity: 75.0%; BMI: 81.3%), RD (obesity: 75.0%; BMI: 77.1%), and AD (obesity: 68.8%; BMI: 70.8%).

**Figure 3:**
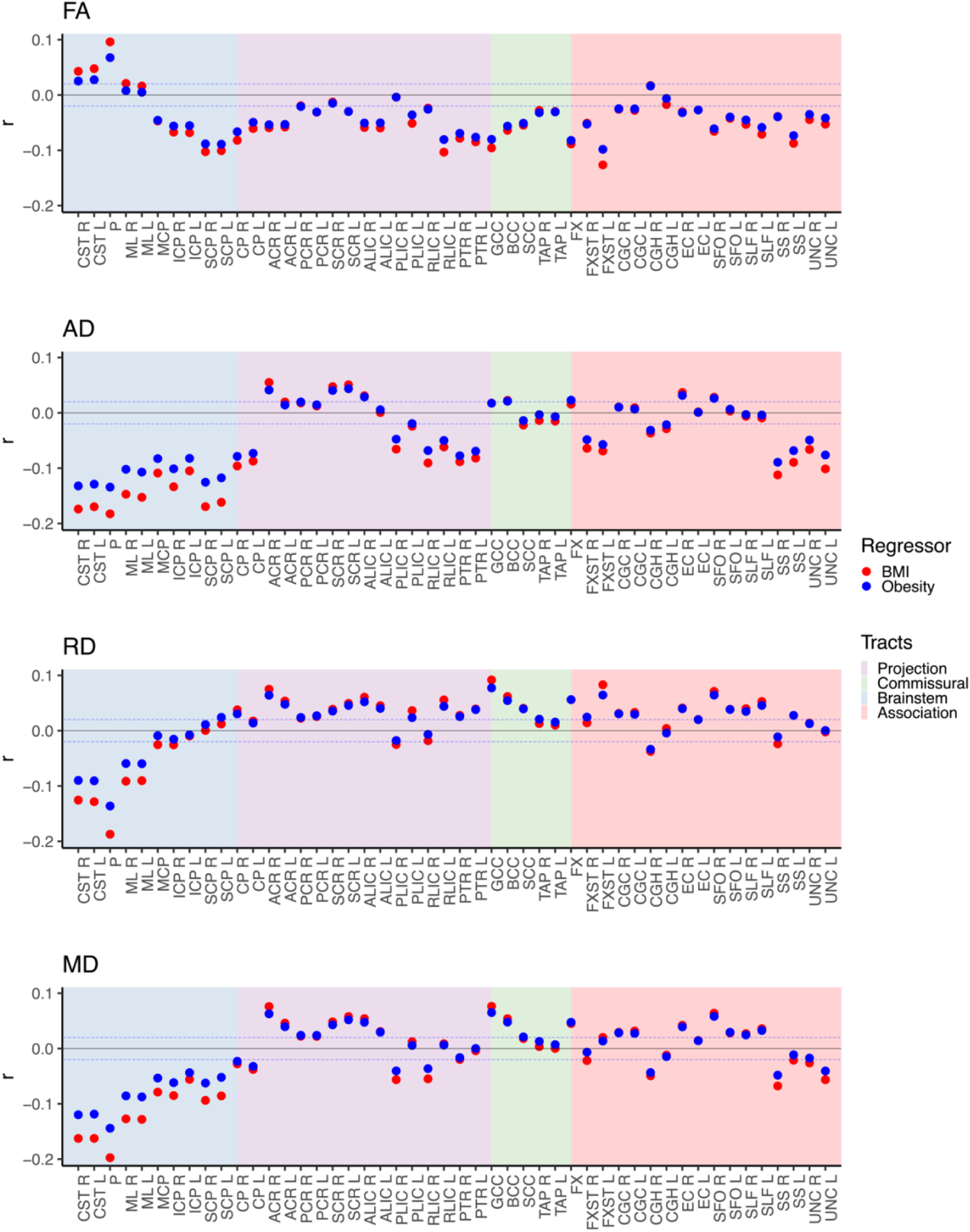
Obesity-related traits with DTI metrics. The figure shows the multiple linear regression results of (i) obesity vs non-obesity and (ii) BMI separately on DTI metrics after adjusting for age, age^2^, sex, age-by-sex, age^2^-by-sex, ethnicity, and site. The blue dotted lines indicate r=±0.02 (corresponds to r effects approximately at significance threshold p≤7.7e-05). Abbreviations: BMI – body mass index; FA – fractional anisotropy; AD – axial diffusivity; RD – radial diffusivity; MD – mean diffusivity; L – left; R – right; r - partial correlation coefficient. For regional white matter abbreviations, see Figure 2.

We observed the overall most significant effects with *brainstem tracts* for both obesity and BMI across DTI metrics, with mixed effect directionality for FA, and predominantly lower AD, RD, and MD effects (**Figure 3**). For FA, the most significant associations were with higher pontine and lower superior cerebellar peduncle effects. The corticospinal tract also exhibited higher FA effects, while the inferior and middle cerebellar peduncle exhibited lower FA effects. FA of the right medial lemniscus was significant for BMI only. Additionally, we further observed the overall lowest AD and RD effects for the pontine followed by the corticospinal tract – the same tracts with the significantly higher FA effects – and the superior cerebellar peduncle (only AD), the tract with the lowest FA effect. MD showed the overall lowest effects for the pontine, followed by the corticospinal tract and medial lemniscus (which only showed significant FA for BMI and the right medial lemniscus) before the superior cerebellar peduncle. These findings suggest that both obesity and higher BMI are associated with lower directional and overall diffusion for brainstem tracts, as well as lower perpendicular diffusion for the corticospinal tract, pontine, and medial lemniscus – the tracts with significantly higher or non-significant FA effects.

For *projection*, *commissural*, and *association pathways*, both obesity and BMI were associated with predominantly lower FA effects, together with lower AD and higher RD and MD effects (although several ROIs also show opposing effect directions; **Figure 3**). These findings indicate that various tract properties contribute to lower FA (e.g., predominantly lower AD or higher RD, or in combination) for both obesity and BMI. The effect sizes were notably smaller than for brainstem tracts, and the patterns not as clear, making it more challenging to distinguish generalizable patterns for non-brainstem tracts.

WHR and waist circumference showed similar associations with DTI metrics (**Figure S3**; **Tables S4-S5**) with a slightly higher number of significant ROIs (82.3% WHR and 82.8% waist circumference) than obesity and BMI, possibly reflecting larger sensitivity to abdominal fat. The significant effect size ranges were for WHR |*r*| in [0.02, 0.13] (p in [1.3e-05, 2.1e-142]) and for waist circumference |*r*| in [0.02, 0.17] (p in [7.5e-05, 1.5e-246]). Thus, the effect sizes were overall lower than for BMI, while the range for obesity fell between that of WHR and waist circumference.

### Interaction effects between sex and obesity-related traits on DTI metrics

Sex by obesity-related traits analyses (**Step 2**) revealed significant interaction effects for a substantial number of ROIs across DTI metrics (obesity: |*r*| in [0.02, 0.05], p in [6.6e-05, 1.4e-27]; BMI: |*r*| in [0.02, 0.06], p in [6.8e-05, 2.5e-32]; **Figure 4**; **Tables S6-S7**). The number of significant sex interactions effects varied across DTI metrics, with higher proportion for AD (obesity: 50.0%; BMI: 41.7%), RD (obesity: 27.1%; BMI: 58.3%), and MD (obesity: 60.4%; BMI: 58.3%) than FA (obesity: 4.2%; BMI: 18.8%).

**Figure 4:**
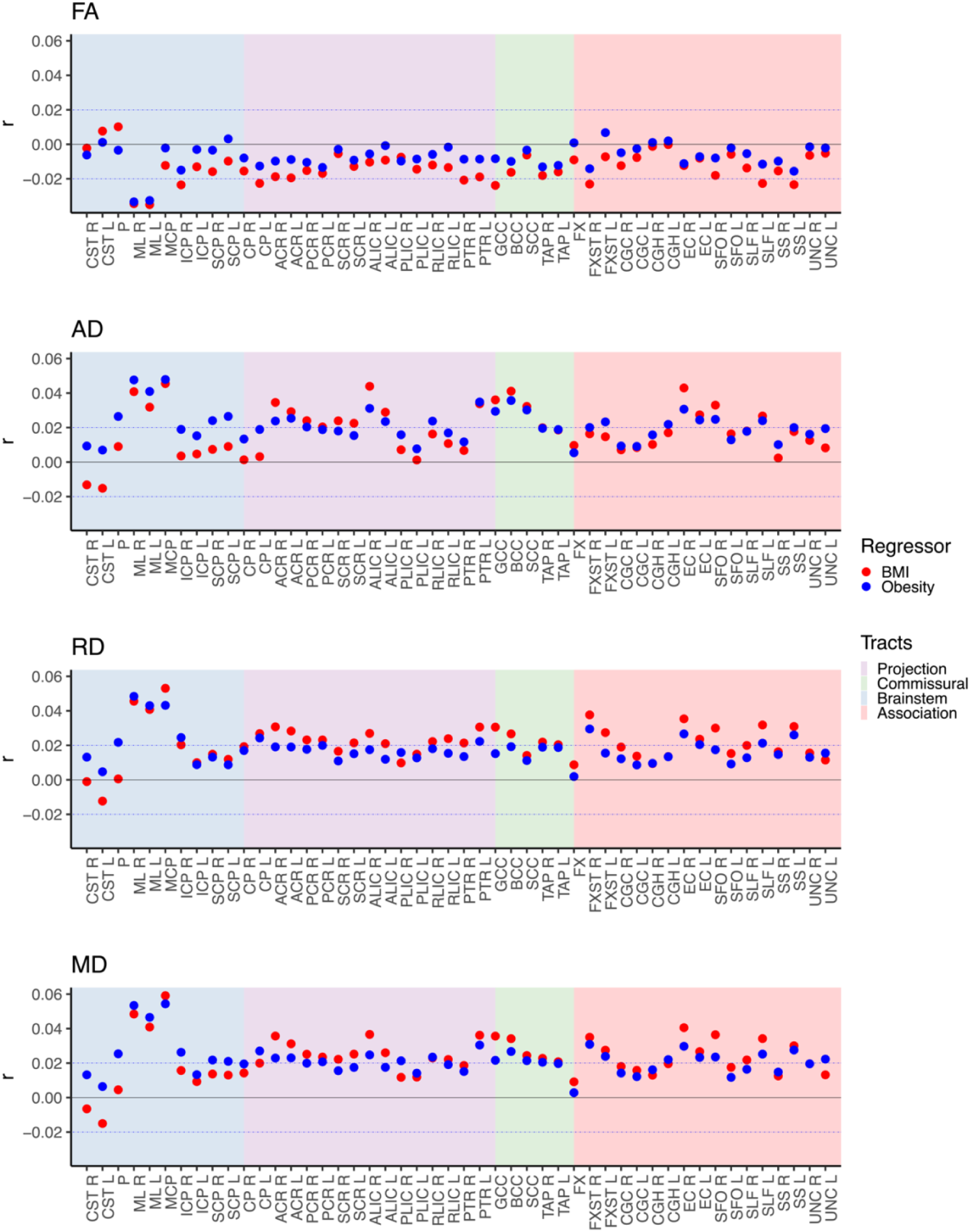
Interaction effects between sex and obesity-related traits on DTI metrics. The figure shows the interaction effects between sex (female reference) and (i) obesity vs non-obesity and (ii) BMI separately on DTI metrics after adjusting for the corresponding main effects, age, age^2^, ethnicity, and site. The blue dotted lines indicate r=±0.02 (corresponds to r effects approximately at significance threshold p≤7.7e-05). Abbreviations: BMI – body mass index; FA – fractional anisotropy; AD – axial diffusivity; RD – radial diffusivity; MD – mean diffusivity; L – left; R – right; r - partial correlation coefficient. For regional white matter abbreviations, see Figure 2.

The findings indicate regional sex-related white matter variation of obesity-related traits. We observed the overall largest interaction effects for two brainstem tracts, the medial lemniscus (FA, AD, RD, and MD) and the middle cerebellar peduncle (AD, RD, and MD only). Therefore, we further illustrated the interaction effects of sex-by-BMI for these two tracts (**Figure 5A-C**). The illustrations show that for the left and right medial lemniscus, the higher FA and lower AD, RD, and MD effects at higher BMI are attenuated in males relative to females. For the middle cerebellar peduncle, we observed a similar pattern for AD and MD, suggesting attenuated reduction at higher BMI in males relative to females, while we observed opposing effects for RD.

**Figure 5:**
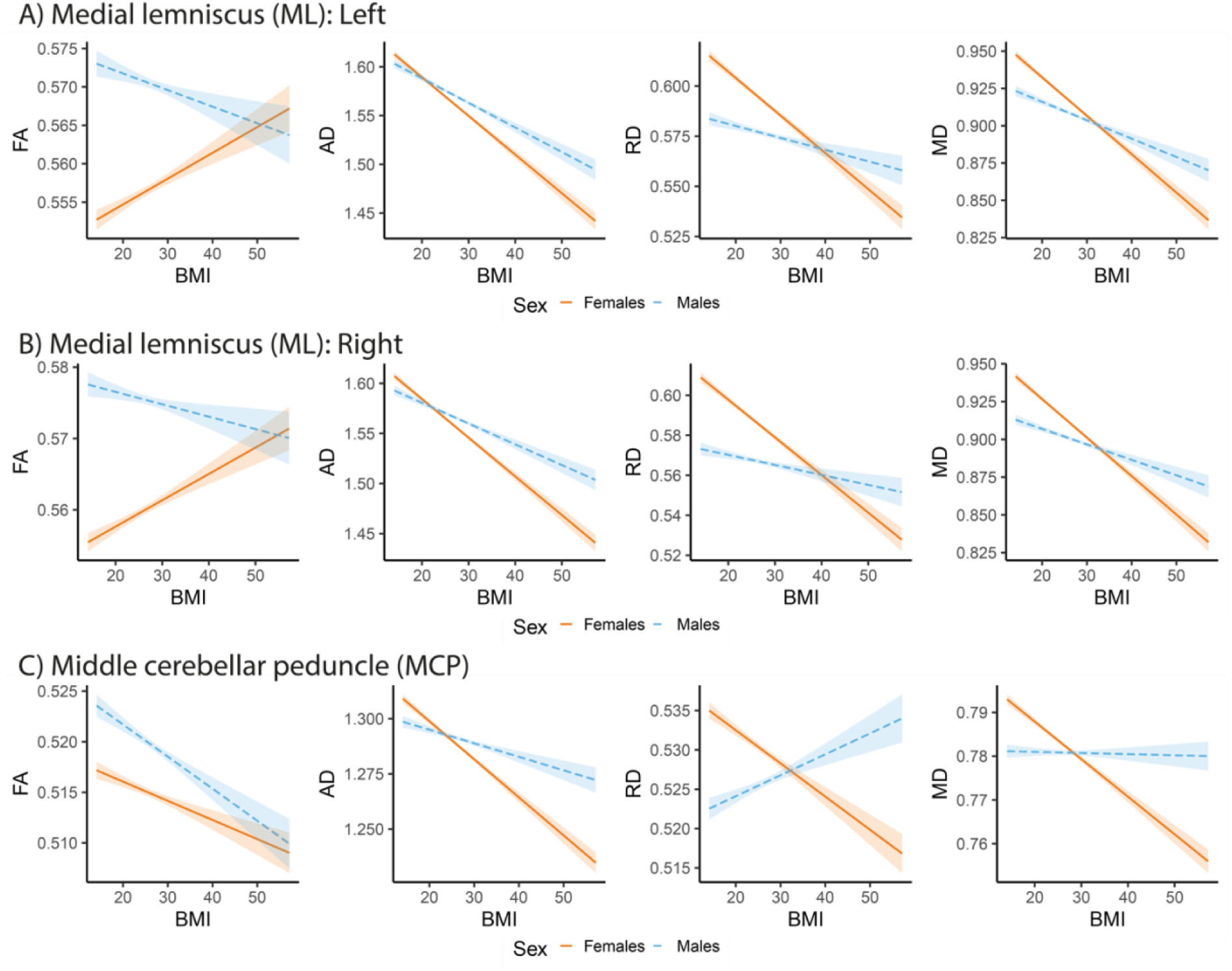
Illustration of interaction effects between sex and BMI for selected regions of interests. Sex-by-BMI interaction effects of the (A) left ML, (B) right ML, and (C) MCP using the interact_plot R-function after adjusting for the corresponding main effects, age, age^2^, ethnicity, and site (BMI uncentered) and 95% confidence intervals. The figures show the effects/fitted lines with confidence/predicted intervals. Abbreviations: BMI – body mass index; FA – fractional anisotropy; AD – axial diffusivity; RD – radial diffusivity; MD – mean diffusivity; L – left; R – right; r - partial correlation coefficient.

Additionally, we observed significant interactions effects between WHR and waist circumference, albeit with some regional differences (**Figure S4**; **Tables S8-S9**).

### Interaction effects between Age and obesity-related traits on white matter microstructure

Lastly (**Step 3**), the age by obesity-related traits analyses revealed significant interactions for some ROIs across DTI metrics (obesity: 2.6%, |*r*| in [0.02, 0.03], p in [2.3e-05, 4.5e-09]; BMI: 4.2%, |*r*| in [0.02, 0.04], p in [6.9e-05, 5.4e-14]). We observed the largest number of significant ROIs for AD (obesity: 6.3%; BMI: 8.3%), followed by MD (obesity: 4.2%; BMI: 2.1%), FA (obesity: 0%; BMI: 4.2%), and RD (obesity: 0%; BMI: 2.1%; **Figure S5**; **Tables S10-S11**).

The analysis further revealed a higher number of significant ROIs for WHR and waist circumference with stronger and more widespread interaction effects with age particularly for WHR (WHR: 27.6%, |*r*| in [0.02, 0.05], p in [7.6e-05, 7.0e-22]; waist circumference: 15.6%, |*r*| in [0.02, 0.03], p in [7.6e-05, 1.0e-11]; **Figure 6**; **Tables S12-S13**) than observed for obesity and BMI, possibly reflecting greater sensitivity to abdominal obesity for both WHR and waist circumference. The largest number of significant ROIs were found for AD (WHR: 39.6%; waist circumference: 31.4%), followed by MD (WHR: 27.1%; waist circumference: 16.7%), FA (WHR: 25.0%; waist circumference: 8.3%), and RD (WHR: 18.8%; waist circumference: 6.6%).

**Figure 6:**
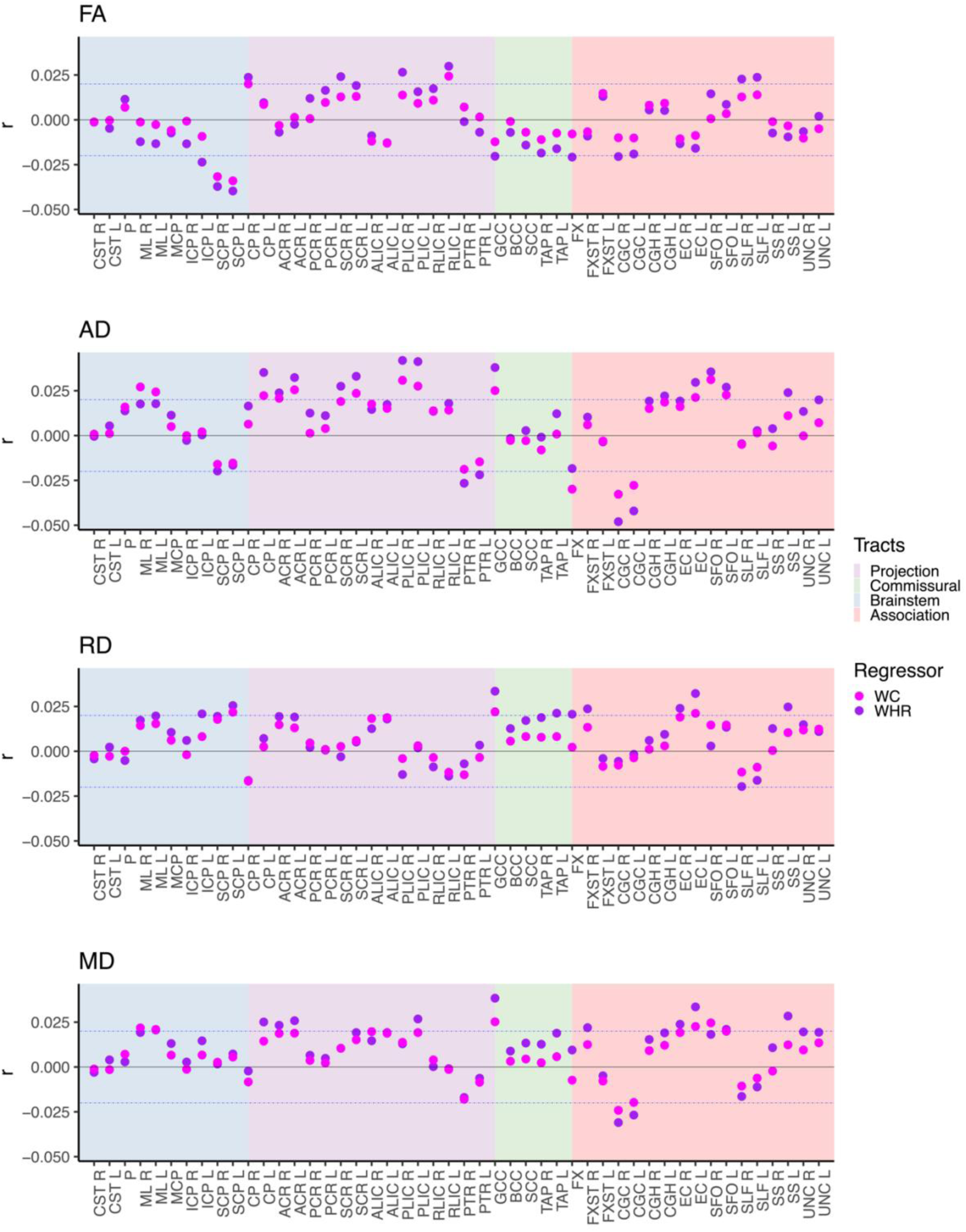
Interaction effects between age and obesity-related traits on DTI metrics. The figure shows the interaction effects between age and (i) WHR and (ii) waist circumference on DTI metrics after adjusting for the corresponding main effects, and sex, ethnicity, and site. The blue dotted lines indicate r=±0.02 (corresponds to r effects approximately at significance threshold p≤7.7e-05). Abbreviations: FA – fractional anisotropy; AD – axial diffusivity; RD – radial diffusivity; MD – mean diffusivity; L – left; R – right; r - partial correlation coefficient; WHR – waist-to-hip ratio; WC – waist circumference. For regional white matter abbreviations, see Figure 2.

In the following, we focused on the tracts with the most significant findings limited to WHR due to the largest number of significant interactions with age on white matter (**Figure 6**; **Tables S12**). We observed the overall highest interaction effects between WHR and age for AD of the *posterior limb of the internal capsule* and AD and MD of the *genu of corpus callosum*, and the overall lowest interaction effect sizes for the AD of the *cingulum cingulate gyrus* and FA of the *superior cerebellar peduncle*. The interpretation of the results varies for the various tracts – all depending on the main effect of WHR. For the *posterior limb of the internal capsule*, there is a significant interaction effect between age and WHR despite no main effect of WHR (**Figure 7A**). Contrastingly, for the *genu of the corpus callosum*, the main effect of WHR is substantial, and the significant interaction with age suggests steeper age-related alterations at higher WHR at higher ages (across FA, AD, RD, and MD) (**Figure 7B**). For the *cingulum*, the main effect of WHR was attenuated by the negative interaction with age and became weaker at higher ages (**Figure 7C**). Like the genu of the corpus callosum, the negative main effect of the *superior cerebellar peduncle* (FA and AD) was substantial, and the results further indicate steeper alteration at higher WHR and higher ages (**Figure 7D**).

**Figure 7:**
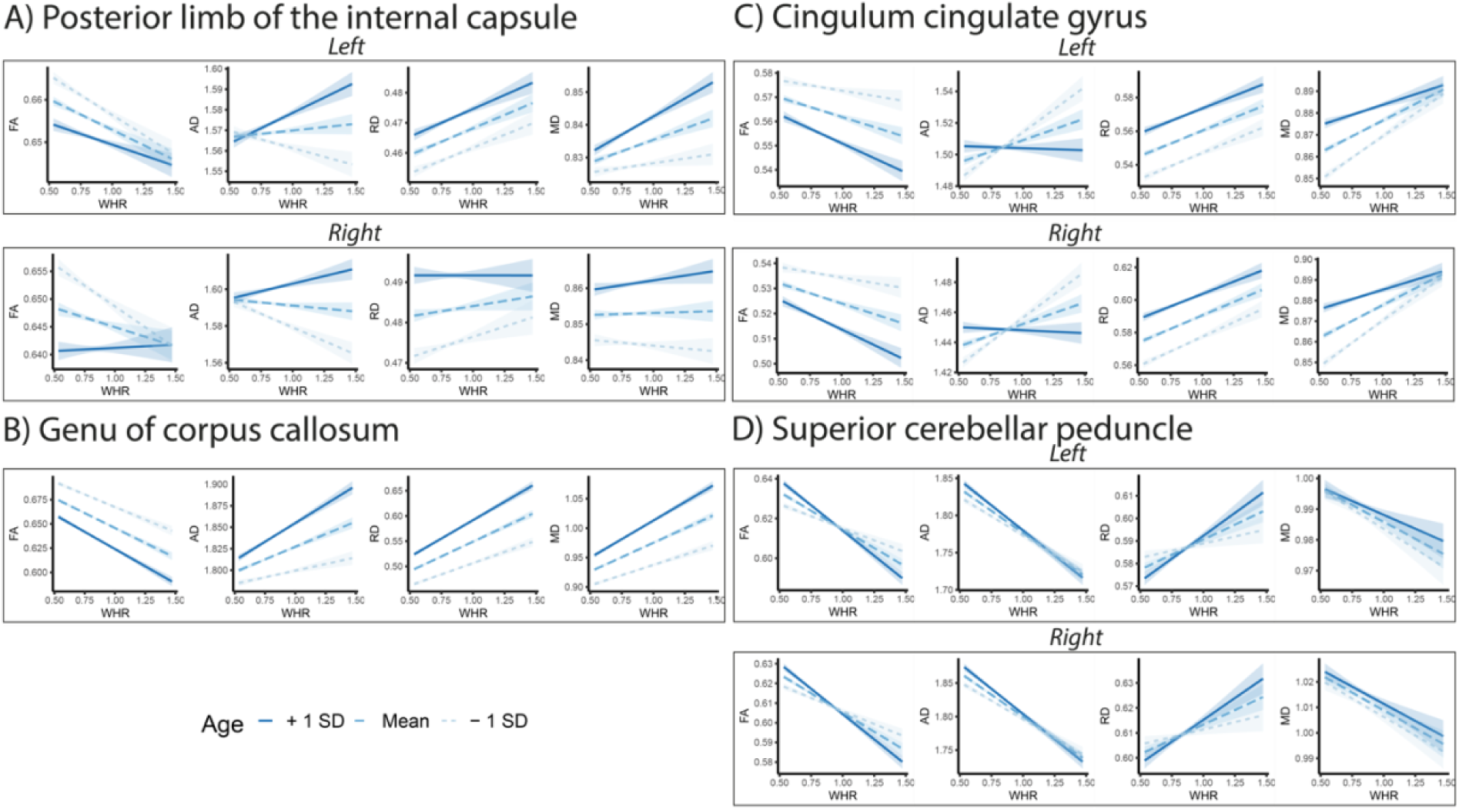
Illustration of WHR by age interaction on white matter ROIs. (**A-D**) shows the interaction effects between WHR and age on selected ROIs using the interact_plot r-function (WHR and age uncentered, with 95% confidence intervals). The statistical model was adjusted for the main effect of WHR and age, sex, ethnicity, and site. Abbreviations: FA – fractional anisotropy; AD – axial diffusivity; RD – radial diffusivity; MD – mean diffusivity; L – left; R – right; r - partial correlation coefficient; WHR – waist-to-hip ratio; WC – waist circumference.

## Discussion

With this study we aimed to elucidate whether and how obesity-related traits relate to regional brain white matter phenotypes, as well as putative sex- and age-related differences. We illustrated that there are (i) widespread obesity-related associations with regional brain white matter phenotypes across DTI metrics, with (ii) the largest effects for brainstem tracts; and that there are (iii) sex- and age-interactions with obesity-related traits on white matter phenotypes, with (iv) more widespread sex-than age-related effects. These findings support a negative link between higher measures of obesity-related traits and widespread regional white matter microstructure alterations with largest effects for the brainstem that may differ based on sex and age. In turn, these findings may relate to the observed comorbid link between cardiometabolic conditions and brain disorders, as well as sex- and age-related variations in brain disorders, and may imply interactive metabolic processes and body fat-related pathways between physical and brain health.

Our most striking finding is the relatively strong associations between obesity-related traits and brainstem tracts of generally lower AD, RD, and MD, which may culminate in either lower or higher FA at higher obesity-related traits, all depending on the underlying diffusion properties. The revealed patterns are in line with a prior study using a partly overlapping sample^4^, and further suggests axonal degeneration with little indications of demyelination and inflammatory processes (as indicated by lower AD, RD, and MD)^16^ in the brainstem in obesity, irrespective of whether FA is higher or lower. Brainstem tracts connect the spinal cord, brainstem, cerebellum, subcortical structures (e.g., thalamus), and/or cerebrum^35^. Although speculative, the observed patterns may implicate the brainstem body-brain communication hub in obesity. Indeed, they may relate to impairments in the brainstem’s role in regulating appetite and food intake in obesity, given that nutritional information is passed from the gut via vagal sensory nerves to the brainstem^36^ and hormones produced in the gut are linked to nutrient absorption and metabolism and appetite regulation in the brain^36^. Another possible explanation is the involvement of the sensorimotor system, where a more sedentary lifestyle in obesity may result in less maintenance or development, or more degeneration, of the important brainstem body-brain communication pathways. Indeed, we have previously shown brainstem and cerebellum volumes alterations for sarcopenic^18^ and obesity-related^37^ traits in partly overlapping samples. Furthermore, structures of the sensorimotor system - including the brainstem and cerebellum - mediated the link between sarcopenic traits and lower cognitive performance^18^, and similar patterns may explain the observed link between midlife obesity and later cognitive deficits^38^. Notably, however, these structures are often not included in large-scale studies of brain disorders, with some exceptions (e.g., corticospinal tract or in studies of ataxia)^10^. Given the overrepresentation of cardiometabolic comorbidities in brain disorders^1–3^ – suggestive of a link between physical and brain health – it is paradoxical that structures connecting the body with the brain, and vice versa, are often not included in larger studies. Thus, future large-scale studies of brain disorders that target structures interconnecting the body with the brain, and vice versa, are warranted to enhance our understanding of brain disorders and the physical, cardiometabolic, link.

We also found that obesity-related traits are associated with widespread regional white matter phenotypes across DTI metrics. Indeed, the findings suggests that the obesity connection with white matter is more systemic than localized, with widespread associations, albeit with the strongest effects for brainstem tracts. The direction of effects varied by region, but for non-brainstem tracts the predominant picture is of lower FA in combination with predominantly lower AD and higher RD and MD (albeit with some mixed effects for AD and MD), which may indicate a pattern of regional axonal degeneration, demyelination, and inflammatory processes (as indicated by lower AD, and higher RD and MD)^16^ of non-brainstem tracts in obesity. These findings align with prior studies showing lower FA at higher BMI or WHR^4–8^ and with observations from smaller studies suggesting axonal degeneration and demyelination for obesity-related traits^5^. However, to our knowledge, no prior large-scale study has comprehensively shown indications of brain axonal degeneration, demyelination, and inflammatory processes in obesity for non-brainstem tracts.

Furthermore, our results show that sex is an essential factor that needs consideration when addressing obesity-related traits and white matter microstructure, which may relate to sex differences in healthy body fat percentage and body fat distributions, as well as hormonal status. The range of normal body fat is higher for females (15-30%) than for males (10-20%)^27^. Females also naturally accumulate more body fat across the hips and thighs, while males often accumulate body fat across the trunk and abdomen^11^, which in turn may be influenced by hormonal status^39^. Indeed, males generally have higher levels of visceral fat than females, although menopausal and post-menopausal females may shift to accumulate body fat more similarly to males^11^. This may suggest different body fat-related aging trajectories in males and females, with the additional complexity of menopause for females. Thus, understanding the observed sex differences in how obesity-related traits relate to white matter microstructure might be important for disentangling sex-specific physical and brain disease risks and outcomes.

We also found regional age-by-obesity-related interactions on white matter microstructure, with larger effects for measures sensitive to abdominal obesity. This may relate to age-related increases in abdominal obesity particularly for males and postmenopausal females^11^, suggestive of sex-specific aging trajectories that might also be reflected in brain structure and possibly in disorders. Such interaction effects between predominantly abdominal obesity-related traits and age on white matter microstructure may form the foundation for future works, that also targets putative sex-specific aging trajectories, the added complexity of menopause in females, and links to health outcomes in aging.

Taken together, our findings support that cardiometabolic factors are of importance for brain health. However, we do not know the causal direction of effects, and indications of reciprocal complex mechanisms exist. Cardiometabolic factors, heart phenotypes, and cardiometabolic multimorbidity are associated with white matter phenotypes, cognitive functioning, and several brain disorders^1,2,40–43^. Similarly, several brain disorders are associated with cardiometabolic factors, cognitive functioning, and white matter phenotypes^1,2,10,42,44,45^. Danish population registry studies further suggest that most categories of physical conditions show higher risk for developing mental disorders^46^, and likewise that most mental disorders are associated with higher risk for developing physical conditions^47^. Additionally, we have illustrated that brain white matter phenotypes mediates the link between sarcopenic-traits^18^ or accumulated liver fat^43^ and lower cognitive performance, and a recent review suggested elevated prevalence of physical conditions in major depression disorder, and vice versa^2^. These observations suggest interconnected and reciprocal links between the body and the brain that may involve common body fat-related pathways, and may indicate that a whole-body approach and novel treatment strategies capturing the body-brain link (e.g., possibly GLP-1^48^ or similar drugs) may be beneficial for treating comorbid brain and cardiometabolic and other physical conditions.

This study offers notable strengths. We included a well-characterized sample of unprecedented size. We performed analyses across commonly used conventional DTI metrics, modeled sex and age specific effects, and illustrated that several DTI metrics are needed to disentangle the underlying tissue properties. The study addresses gaps in the scientific literature and provides novel insight into the complexity linking sex, age, and obesity-related traits with white matter microstructure, which may be relevant for health and disease trajectories. The study also had some limitations. The sample is healthier than the general population^49^ and predominantly includes self-identified white Europeans. It was beyond the study’s scope to investigate specific clinical conditions, other cardiometabolic risk factors, different body fat compartments, metabolically healthy and unhealthy obesity^27^, or putative shared genetic factors^2,41^ with white matter phenotypes, which may provide added and more specific information. We focused our analyses on the conventional DTI model^15^, but other dMRI analysis models (e.g., restriction spectrum imaging^50^, neurite orientation dispersion and density imaging^51^) may provide complementary information^52^. We used multiple linear regression for the statistical analyses, but future work may consider using multimodal techniques that reduce the dimensionality of the data and facilitate interpretation^53^. We focused on the most significant findings and overall patterns, but the vast number of significant findings suggests that future studies are needed to disentangle the regional complex interplay of sex, age, and obesity-related traits on white matter, and putative links to disease onset, progression, and outcome.

To conclude, we observe that obesity-related traits are associated with widespread white matter microstructure phenotypes, with the largest effects for brainstem tracts; a key hub interconnecting the brain with the rest of the body (and vice versa) that is involved in appetite regulation. Furthermore, our analysis indicated more widespread sex-than age-related interaction effects with obesity-related traits on white matter phenotypes in this sample. Our findings support a link between cardiometabolic and brain health and suggest that addressing obesity-related risk factors may be important for prevention, treatment, and outcome of comorbid physical and brain disorders, although further studies are needed to examine causal effects.

## Acknowledgments

This research has been conducted using the UK Biobank Resource under Application Number 27412 and uses data provided by patients and collected by the NHS as part of their care and support. We performed all data analyses on the *Services for Sensitive Data* (TSD), University of Oslo, Norway, with resources provided by UNINETT Sigma2 - the National Infrastructure for High-Performance Computing and Data Storage in Norway.

## Data availability

The UK Biobank resource is open for eligible researchers upon application (http://www.ukbiobank.ac.uk/register-apply/).

## Code availability

We used publicly available resources to process the brain image data and conduct statistical analyses. We extracted data from individual UK biobank baskets using the *ukb_helper.py* script (https://github.com/precimed/ukb). The project R-scripts are publicly available at: https://osf.iodoi (upon publication).

## Authors’ contributions

*Study design:* TPG, DB, IV, LTW, OAA. *Data preparation and image processing:* TPG, IM. *Data management:* TPG, IM. *Analytical strategy:* TPG, DB, IV, LTW, OAA. *Statistical analysis:* TPG. *Figures:* TPG. *Data* i*nterpretation*: TPG, DB, NP, IV, LTW, OAA. *Manuscript preparation:* TPG, DB, IV, LTW, OAA. *Funding:* TPG, OAA, LTW. All authors revised the manuscript and approved the final version.

