## Supplementary information for "Mapping obesity-related traits with regional white matter microstructure highlights the importance of brainstem tracts and sex-related differences"

### SUPPLEMENTAL TABLES

**TABLE S1: DEMOGRAPHIC AND CLINICAL DATA.**

| | Men vs. women | | | | Participants with obesity vs. non-obesity | | | |
|---|---|---|---|---|---|---|---|---|
| | **Men (N=19,136)** | **Women (N=20,904)** | **test** | **p-value** | **Obesity (N=6,912)** | **Non-obesity (N=33,128)** | **Test** | **p-value** |
| **Women** | | | | | 3507 (50.7) | 17397 (52.5) | 7.2 | 0.0074 |
| **Age (year)**[1] | 64.9±7.8 | 63.7±7.6 | 15.7 | <0.0001 | 63.5±7.6 | 64.4±7.8 | −8.9 | <0.0001 |
| **Age range (year)** | [44 83] | [45 83] | | | [44 83] | [45 83] | | |
| **European ethnicity**[2] | 18563 (97) | 20293 (97.1) | 0.2 | 0.6949 | 6697 (96.9) | 32159 (97.1) | 0.6 | 0.4300 |
| **Cigarette smoker**[2] | 601 (3.1) | 470 (2.2) | 30.2 | <0.0001 | 192 (2.8) | 879 (2.7) | 0.3 | 0.5877 |
| **Smoker current / previous / never** | 601 / 7049 / 11486 | 470 / 6432 / 14002 | | | 192 / 2626 / 4094 | 879 / 10855 / 21394 | | |
| **Alcohol consumer**[2] | 18125 (94.7) | 19380 (92.7) | 67.5 | <0.0001 | 6394 (92.5) | 31111 (93.9) | 18.8 | <0.0001 |
| **Alcohol consumer current / previous / never** | 18125 / 587 / 424 | 19380 / 679 / 845 | | | 6394 / 247 / 271 | 31111 / 1019 / 998 | | |
| **Obesity**[2] | 3405 (17.8) | 3507 (16.8) | 7.2 | 0.0074 | | | | |
| **Height (cm)**[1] | 176±6.6 | 162.7±6.2 | 207 | <0.0001 | 168.6±9.4 | 169.2±9.2 | −4.6 | <0.0001 |
| **Weight (kg)**[1] | 83.3±13.2 | 68.7±12.9 | 112.1 | <0.0001 | 95.3±13.5 | 71.6±11.6 | 135.6 | <0.0001 |
| **BMI**[1] | 26.9±3.8 | 25.9±4.7 | 21.7 | <0.0001 | 33.4±3.4 | 24.9±2.7 | 197 | <0.0001 |
| **Waist circ. (cm)**[1,3] | 94.1±10.5 | 82.6±11.7 | 102.7 | <0.0001 | 103.9±10.4 | 84.8±10.3 | 140.1 | <0.0001 |
| **Hip circ. (cm)**[1] | 100.5±7.3 | 100.6±9.8 | −1.8 | 0.0773 | 112.4±8.6 | 98.1±6.3 | 131.4 | <0.0001 |
| **WHR**[1] | 0.9±0.1 | 0.8±0.1 | 172.6 | <0.0001 | 0.93±0.09 | 0.86±0.08 | 52.7 | <0.0001 |
| **Diabetic**[2] | 469 (2.5) | 253 (1.2) | 86.1 | <0.0001 | 291 (4.2) | 431 (1.3) | 271.7 | <0.0001 |
| **High cholesterol**[2] | 3195 (16.7) | 2008 (9.6) | 443.6 | <0.0001 | 1209 (17.5) | 3994 (12.1) | 148.9 | <0.0001 |
| **Hypertension**[2] | 4768 (24.9) | 3500 (16.7) | 406.8 | <0.0001 | 2371 (34.3) | 5897 (17.8) | 949.4 | <0.0001 |

*Notes:* The table reports mean ± standard deviation for continuous variables, and N (%) for categorical variables. Footnotes describe the applied test. Obesity is defined as BMI ≥30, and self-reported data was used to determine ethnicity, diagnosis of diabetes, high cholesterol, and hypertension, and current alcohol consumption and cigarette smoking. *Abbreviations:* BMI – body mass index; circ. – circumference; WHR – waist-to-hip ratio.

---

[1] Welch Two Sample t-test

[2] Pearson's Chi-squared test with Yates' continuity correction

[3] Two sample t-test (Participants with obesity vs. non-obesity only)

### SUPPLEMENTAL FIGURES

**FIGURE S1: FLOWCHART - OVERVIEW OF THE PARTICIPANT INCLUSION PIPELINE.**

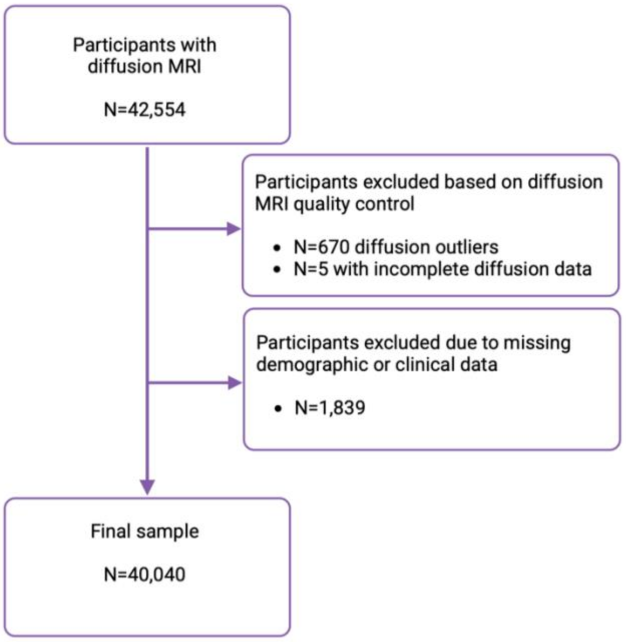

Notes: Created with BioRender.com

**FIGURE S2: DENSITY PLOTS OF THE INCLUDED CONTINUOUS VARIABLES: IMAGING ASSESSMENT.**

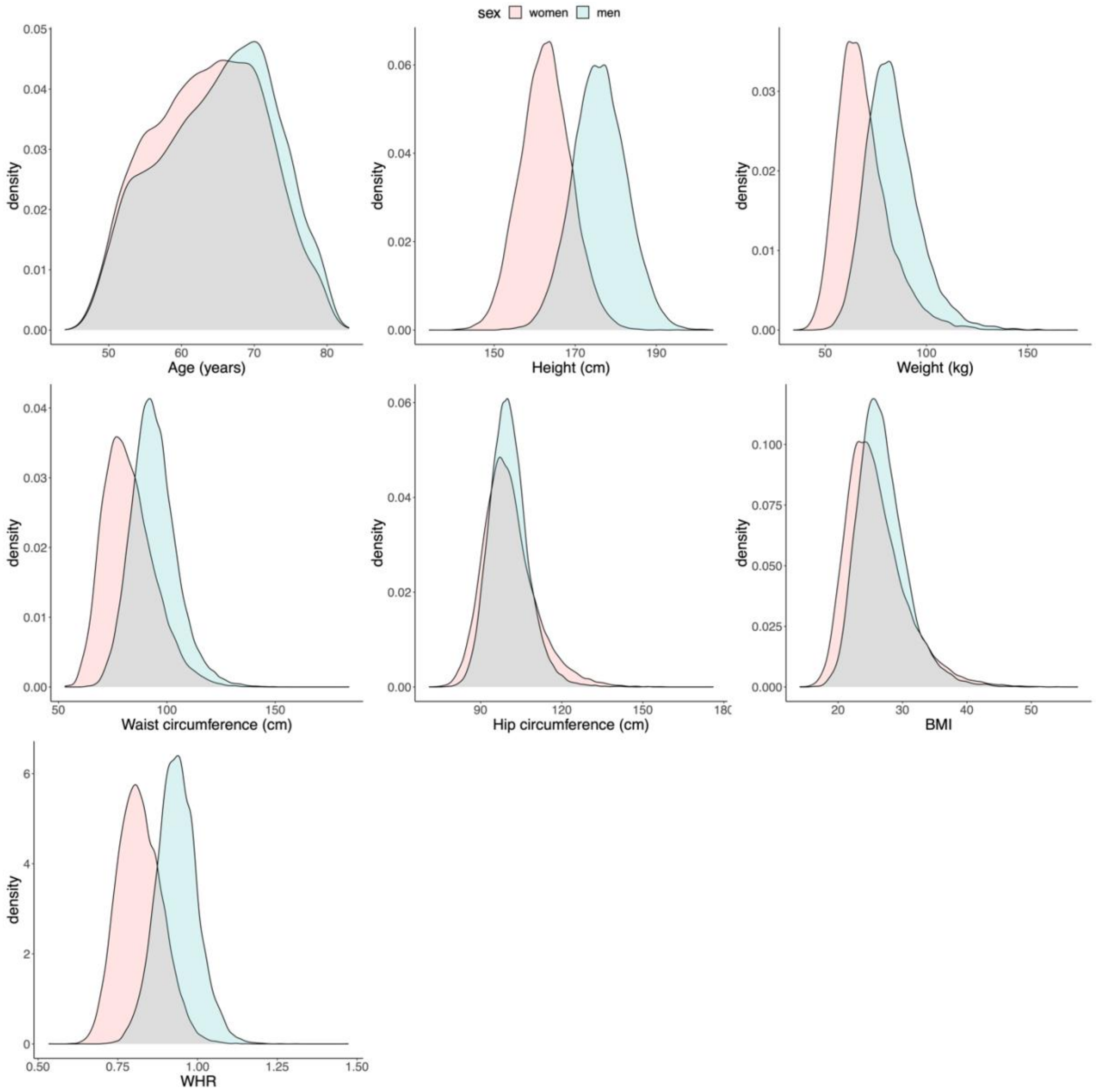

Notes: Density plots split on sex from the imaging timepoint. Abbreviations: BMI – body mass index; WHR – waist-to-hip ratio.

**FIGURE S3 THE PATTERNS OF OBESITY-RELATED BODY TRAITS ON WHITE MATTER MICROSTRUCTURE**

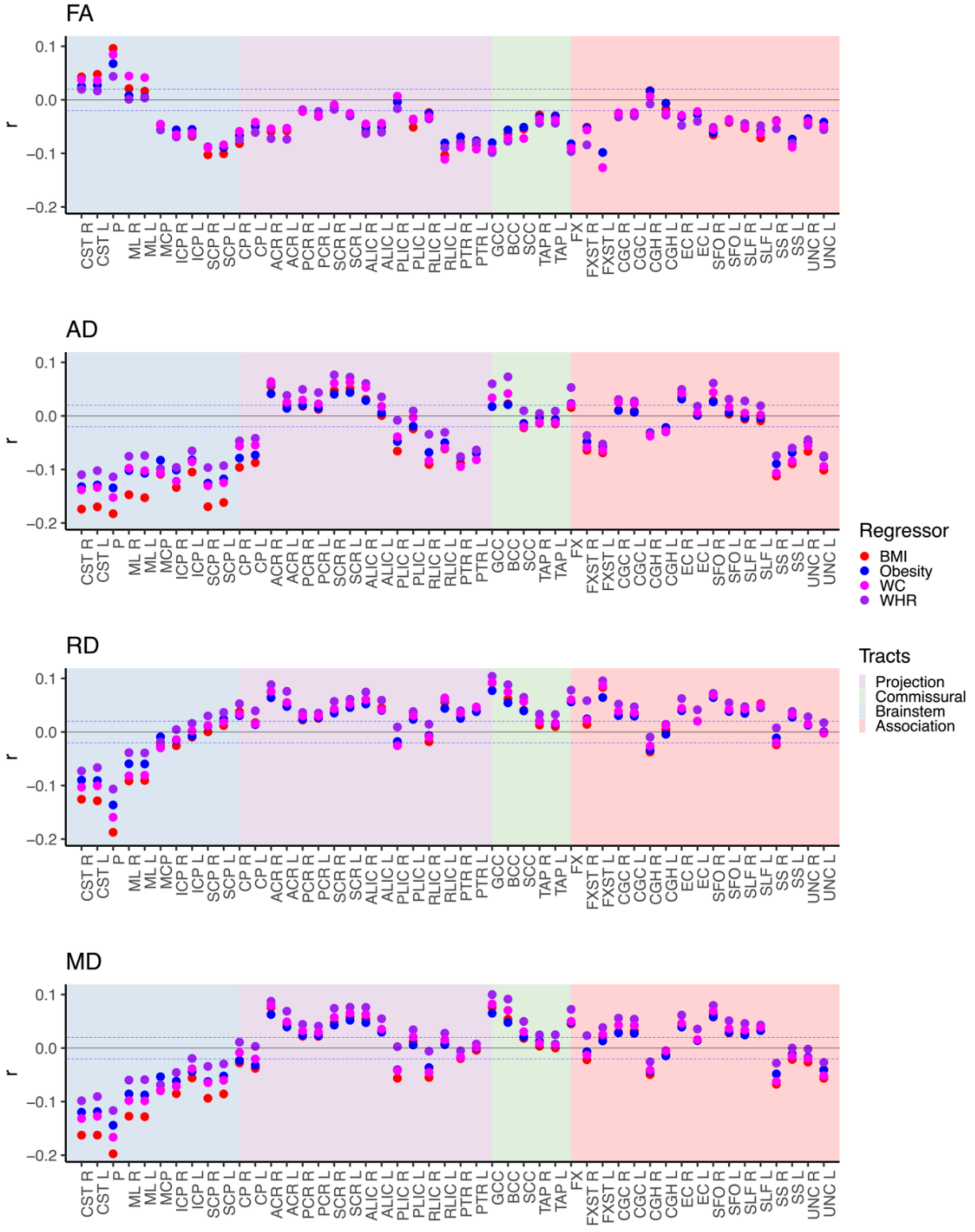

*Notes:* The figure shows the multiple linear regression results of obesity-related traits (obesity vs non-obesity, BMI, WHR, and waist circumference) separately on white matter microstructure after adjusting for age, age$^2$, sex, age-by-sex, age$^2$-by-sex, ethnicity, and site. The blue dotted lines indicate r=±0.02 (corresponds to r effects approximately at significance threshold p≤7.7e-05). *Abbreviations*: BMI – body mass index; WC – waist circumference; WHR – waist-to-hip ratio; FA – fractional anisotropy; AD – axial diffusivity; RD – radial diffusivity; MD – mean diffusivity; L – left; R – right; r - partial correlation coefficient; For regional white matter abbreviations, see Figure 2.

**FIGURE S4: SEX-BY-OBESITY-RELATED BODY TRAITS ON WHITE MATTER MICROSTRUCTURE: INTERACTION TERM.**

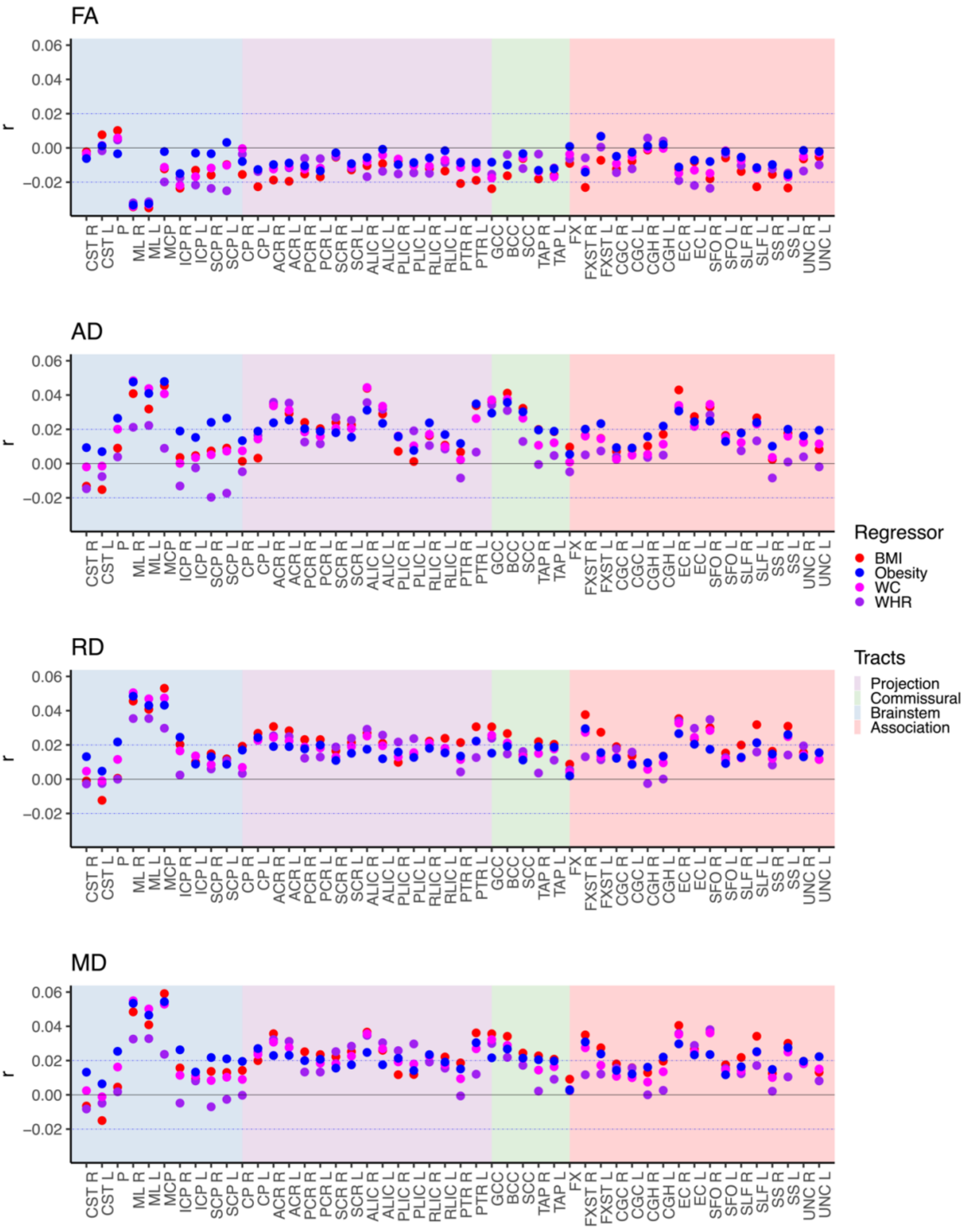

Notes: The figure shows the interaction term of the model investigating sex-by-obesity-related body traits on brain white matter microstructure. We adjusted for the corresponding main effects, age, age2, ethnicity, and site. The blue dotted lines indicate r=±0.02 (corresponds to r effects approximately at significance threshold p≤7.7e-05). Abbreviations: BMI – body mass index; WC – waist circumference; WHR – waist-to-hip ratio; FA - fractional anisotropy; AD – axial diffusivity; RD – radial diffusivity; MD – mean diffusivity; L – left; R – right; For regional white matter abbreviations, see Figure 2.

**FIGURE S5: AGE-BY-OBESITY-RELATED TRAITS ON WHITE MATTER MICROSTRUCTURE: INTERACTION TERM.**

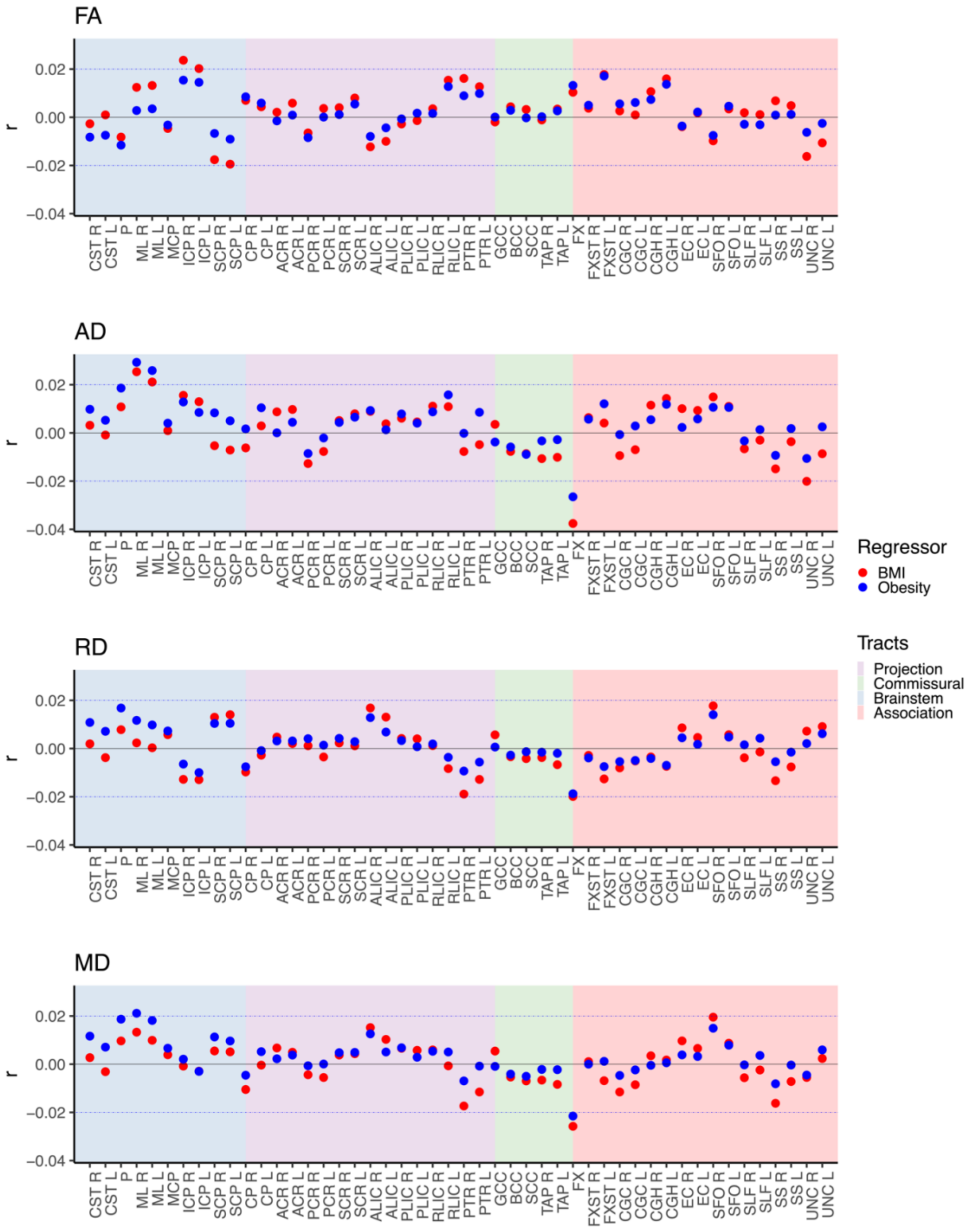

Notes: The figure shows the interaction term of the model investigating age-by-obesity-related trait (here obesity and BMI) interactions on brain white matter microstructure. We adjusted for the main effects, and age, sex, ethnicity, and site. The blue dotted lines indicate r=±0.02 (corresponds to r effects approximately at significance threshold p≤7.7e-05). Abbreviations: BMI – Body mass index; FA - fractional anisotropy; AD – axial diffusivity; RD – radial diffusivity; MD – mean diffusivity; L – left; R – right; For regional white matter abbreviations, see Figure 2.

### Supplemental Notes

#### Note S1: Overview of extracted UK Biobank field IDs.

| Field-ID | Description | Field-ID | Description |
|---|---|---|---|
| 31 | Sex (from central registry, but participants may update it) | 20002 | Self-reported non-cancer diagnosis |
| 54 | Assessment centre | 50 | Standing height (cm) |
| 21003 | Age (years) | 21002 | Weight (kg) |
| 20117 | Alcohol drinker status (current, previous, never) | 21001 | BMI ($kg/m^2$) |
| 20116 | Smoking status (current, previous, never) | 48 | Waist circumference (cm) |
| 21000 | Self-reported ethnic background | 49 | Hip circumference (cm) |

*Notes:* We extracted variables from the imaging time-point when available. For self-reported ethnic background, we complemented missing data with data from the baseline assessment (for details see https://biobank.ndph.ox.ac.uk/showcase/field.cgi?id=21000). Abbreviations: BMI – Body Mass Index.

#### Note S2: Diffusion MRI post-processing.

We post-processed the dMRI data using an optimized post-processing pipeline[1]. Briefly, we corrected for noise using *MP-PCA*[2], Gibbs ringing using *unring*[3], susceptibility distortion using *FSL*[4] *topup*[5], eddy-current and motion-induced distortions using *FSL Eddy*[6–8]. Lastly, we applied isotropic $1mm^3$ Gaussian kernel smoothing to increase the signal-to-noise ratio using *FSL fslmaths*.

Following post-processing, we derived diffusion maps of fractional anisotropy (FA), radial diffusivity (RD), axial diffusivity (AD), and mean diffusivity (MD) using a cumulant expansion of the diffusion signal[9] and MATLAB scripts (https://github.com/NYU-DiffusionMRI/DESIGNER-v1).We normalized diffusion metrics using *tract-based spatial statistics* (TBSS)[10]. We aligned all diffusion maps to the *FSL FMRI58_FA template* using non-linear transformation in *FSL FNIRT*[1]. We derived the mean FA image of all participants, thinned it to create the mean FA skeleton, and projected the scalar diffusion maps onto the FA skeleton. In order to harmonize the obtained diffusion data, we used YTTRIUM algorithm[11]. We extracted the 27 regions of interest (ROIs) for each diffusion map from the Johns Hopkins University (JHU) DTI atlas[12].
